# Socioeconomic inequalities associated with mortality for COVID-19 in Colombia: A cohort nation-wide study

**DOI:** 10.1101/2020.12.14.20248203

**Authors:** Myriam Patricia Cifuentes, Laura A. Rodriguez-Villamizar, Maylen Liseth Rojas-Botero, Carlos Alvarez-Moreno, Julián A. Fernández-Niño

## Abstract

**Background:** After eight months of the COVID-19 pandemic, Latin American countries have some of the highest rates in COVID-19 mortality. Despite being one of the most unequal regions of the world, there is a scarce report of the effect of socioeconomic conditions on COVID-19 mortality in their countries. We aimed to identify the effect of some socioeconomic inequality-related factors on COVID-19 mortality in Colombia.

**Methods:** We conducted a survival analysis in a nation-wide retrospective cohort study of confirmed cases of COVID-19 in Colombia from March 2^nd^ to October 26^th^, 2020. We calculated the time to death or recovery for each confirmed case in the cohort. We used an extended multivariable time-dependent Cox regression model to estimate the hazard risk ratio (HR) by age groups, sex, ethnicity, type of health insurance, area of residence, and socioeconomic strata.

**Results:** There were 1 033 218 confirmed cases and 30 565 deaths for COVID-19 in Colombia between March 2^nd^ and October 26^th^. The risk of dying for COVID-19 among confirmed cases was higher in males (HR=1.68 95% CI: 1.64-1.72), in people older than 60 years (HR=296.58 95% CI: 199.22-441.51), in indigenous people (HR=1.20 95% CI: 1.08-1.33), in people with subsidized health insurance regime (HR=1.89 95% CI: 1.83-1.96), and in people living in the very low socioeconomic strata (HR=1.44 95% CI: 1.24-1.68).

**Conclusion:** Our study provides evidence of socioeconomic inequalities in COVID-19 mortality in terms of age groups, sex, ethnicity, type of health insurance regime, and socioeconomic status.

## INTRODUCTION

The coronavirus disease (COVID-19) is the first pandemic caused by a human coronavirus, the SARS-CoV-2. The first cluster of patients with pneumonia of unknown origin was reported in Wuhan, China in January, 2020.[1] As of October 31^st^, 2020, there were more than 45.5 million confirmed cases and 1.1 million deaths affecting 188 countries around the world. The region of the Americas is the most affected region accounting for more than 20.3 million confirmed cases and 636 482 deaths.[2]

Recently it has been declared that the situation due to COVID-19 corresponds to a syndemic since there is a combination between the epidemic due to the infection by SARS-CoV-2 and the epidemic due to chronic non-communicable diseases that interact in a social context of poverty and inequity.[3] There are three crisis affecting economies and societies in the region: the slow economic growth, the environmental emergency, and the growing inequality.[4] The combination of these social crises with the endemic of non-communicable diseases and the current pandemic are disproportionately affecting the region. Latin America currently holds some of the highest COVID-19 death rates in the world and is facing a humanitarian crisis powered by the longstanding inequality of its countries.[5]

COVID-19 has been recognized by some governments and media as “the great equalizer” due to its capacity to affect people of different age groups, socioeconomic conditions, or prestige.[6] While this is probably true in terms of the biological risk of infection, it is not the case for the observed risk of COVID-19 infection, severity and mortality. There is evidence of racial and socioeconomic disparities in the United States in terms of the population infected by and dying from COVID-19. [7] However, socioeconomic characteristics are not routinely collected or described in most COVID-19 analyses. [8] Therefore, there is a need for collecting and analyzing data on socioeconomic determinants of health to monitor COVID-19 inequities, identify high-risk populations, and guide the development of public health interventions within countries.

During the first wave of the pandemic by SARS-CoV-2 infection/COVID-19 in Colombia, South America, the national public health surveillance system early adapted and prepared for this new threat, being able to detect and follow-up the ongoing cases and their demographic and socioeconomic characteristics. To identify the effect of some demographic and socioeconomic inequality-related factors on COVID-19 mortality during the first eight months of the epidemic in Colombia, we conducted a survival analysis (time to death for COVID-19) using individual data from a nation-wide cohort.

## METHODS

### Study population

Colombia is located in the north corner of South America. According to the National Administrative Department of Statistics (DANE, for its initials in Spanish), the total population is projected by 2020 in 50 372 424 inhabitants.[9] The country is divided into 33 departments and districts which groups 1122 municipalities. Half of the population are women (51.2%), 77.1% of people live in urban areas and 68.2% of Colombians are between 15 and 64 years old. The first case of infection for SARS-CoV-2 was confirmed on March 6^th^ in Bogotá.

### Study design and data sources

We conducted a survival analysis in a nation-wide retrospective cohort study of confirmed cases of COVID-19 in Colombia from March 2^nd^ to October 26^th^, 2020. The nation-wide cohort was ensembled using individual data obtained from the national public health surveillance system (SIVIGILA, for their initials in Spanish). The National Institute of Health (INS, for its initials in Spanish) compiles, verifies and adds laboratory data and other criteria for confirm or discard cases and publishes anonymized and de-identified registries as open data (www.ins.gov.co). The first day of symptoms’ onset for the first confirmed case was February 26^th^ and there were 245 days elapsed till the end of the follow-up period.

Symptomatic and asymptomatic COVID-19 cases are confirmed in Colombia by using Reverse transcription polymerase chain reaction (RT-PCR). Starting on July 23^rd^, 2020, symptomatic cases can be also confirmed by using antigen-based validated tests. Deaths for COVID-19 are notified by health care services to SIVIGILA and DANE and then an individual analysis of the cases confirms, discards or keeps as suspected the reports of deaths due to COVID-19.

All procedures performed in this study followed the national and international ethical standards. Informed consent was not required due to the nature of the study and use of anonymized data from publicly available data sources.

### Outcome and predictors assessment

The outcome event of interest for the study was COVID-19 death among confirmed cases. For deceased symptomatic cases, we computed the ‘time to event’ as the difference between the dates of symptoms onset and the date of death. For deceased asymptomatic cases, ‘time to event’ was calculated as the difference between the date of first medical appointment and the date of death. The follow-up time of symptomatic recovered cases was the difference between the date of symptoms’ onset and date of recovering (registered as laboratory or clinical recovery). For asymptomatic recovered cases, the follow-up time was calculated as the difference between the date of the first medical appointment and the registered date of recovery. We defined censored cases as active cases for which no event (death or recovery) was confirmed by the last date of observation (October 26^th^). For symptomatic and asymptomatic censored cases, we used the same calculation of the follow-up time described above taking into account the date of symptoms’ onset or the date of the first medical appointment date, respectively (See Figure S1 in Supplementary material).

The exposure predictor variables of the model included the following individual demographic and socioeconomic variables that usually leads to health inequalities: age, sex, ethnicity, type of health insurance, area of residence, and socioeconomic strata. In Colombia, ethnicity minorities include “indigenous”, “African-Colombian descent”, a special group of “Raizales” which refers to descendants of the original enslaved Africans, and “Gipsy-Romany”. The type of health insurance is a proxy variable for health access. The “Contributory” type refers to job-related health insurance, the “Subsidized” type covers poor people without formal jobs which hold a subsidy paid by the government, the “Special” health regime covers few unionized workers, and the “exception” regime groups the army-related members. We included age grouped into six life-course categories: infants (0-5y), children and school-age (5-11y), adolescents (12-26y), young adults (27-45y), adults (46-59y), and seniors (60 or more years). The socioeconomic strata is a classification based on the stratification of residential properties used by DANE according to the socioeconomic resources of a census block that received public services. This classification divides houses into class levels, which range from one (very low) to six (high) being one the strata with higher socioeconomic deprivation.[10] The socioeconomic strata is used as a proxy of socioeconomic status (SES) in this study and was obtained as a self-reported variable in SIVIGILA.

### Statistical analysis

All confirmed COVID-19 cases were included in the analysis by using the national cohort’s time to death and recovery. Exploratory customary descriptions for distributions of continuous time-to-death variable and all categorical predictors, for the different outcomes (dead, recovered and censored) included means, medians, frequencies and percentages. The modeling process included testing of proportional hazards assumption for all the predictors using hypothesis tests (p-values of terms addressing time dependent factors) and graphs (Log minus log plots and partial residuals plots from models with no interaction terms). As all predictors but sex were dependent on time, we used an extended multi-predictor time-dependent Cox regression model.[11] By using this extended model it is possible to jointly evaluate the effect of multiple time-dependent variables and their role as potential confounders or effect modifiers. We included simple product interactions between these variables and the time to event to estimate an extended Cox regression model that allows non-proportional hazards. Survival functions were calculated using the Kaplan-Meier method. As our objective was to obtain an explanatory model, we ran the multi-predictor regression models by the *Enter* method, therefore, the resulting equation included all variables. We assessed the coefficient signs and significance by the Wald statistic, and associations expressed as hazard ratios (HRs) with 95% confidence intervals (CIs). All tests with p<0.05 were considered statistically significant. We performed the statistical analysis using SPSS software® version 26.

## RESULTS

There were 1 033 218 confirmed cases and 30 565 deaths for COVID-19 in Colombia from the first day of notification, March 2^nd^, to October 26^th^. Table 1 summarizes the characteristics of the cohort of COVID-19 confirmed cases. Most confirmed COVID-19 cases were male, older than 60 years old, living in urban areas, with the contributory regime of health insurance, and living in residences that belong to the two lower levels of socioeconomic strata. Seven (0.02%) out of the 30 565 cases that end up in deaths were asymptomatic. From all 914 882 confirmed cases that end up in recovery, 11.2% were asymptomatic. The 12.1% of the 87 874 confirmed cases censored at the end of the follow-up time were asymptomatic (See Tables S1-S3 in Supplementary material).

**Table 1.** Sociodemographic characteristics of COVID-19 confirmed cases and deaths in Colombia up to and including October 26^th^, 2020

| Characteristic | Total confirmed cases alive<br>(N= 1 002 653)<br>(n, % for rows) | Total confirmed deaths<br>(N= 30 565)<br>(n, % for rows) | Total confirmed cases<br>(N= 1 033 218)<br>(n, % for columns) |
| --- | --- | --- | --- |
| Sex |  |  |  |
| Male | 500 820 (96.23) | 19 613 (3.77) | 520 433 (50.37) |
| Female | 501 833 (97.86) | 10 952 (2.14) | 512 785 (49.63) |
| Age groups |  |  |  |
| Infants | 20 892 (99.82) | 38 (0.18) | 20 930 (2.03) |
| Children and school-age | 23 391 (99.95) | 12 (0.05) | 23 403 (2.27) |
| Adolescents | 203 475 (99.54) | 231 (0.11) | 203 706 (19.72) |
| Young adults | 430 375 (99.54) | 2000 (0.46) | 432 375 (41.85) |
| Adults | 193 142 (97.48) | 5000 (2.52) | 198 142 (19.18) |
| Seniors | 131 378 (84.95) | 23 284 (15.05) | 154 662 (14.97) |
| Ethnicity |  |  |  |
| White, mestizo, other | 942 712 (97.08) | 28 366 (2.92) | 971 078 (93.99) |
| African-Colombian | 37 883 (96.38) | 1421 (3.62) | 39 304 (3.80) |
| Indigenous | 22 011 (96.59) | 776 (3.41) | 22 787 (2.21) |
| Gipsy-Roman | 34 (94.44) | 2 (5.56) | 36 (0.00) |
| Raizal | 13 (100.00) | 0 (0.00) | 13 (0.00) |
| Area of residence |  |  |  |
| Urban | 861 071 (96.87) | 27 844 (3.13) | 888 915 (86.03) |
| Semirural (village) | 44 586 (97.18) | 1296 (2.82) | 45 882 (4.44) |
| Sparse rural | 25 002 (96.19) | 991 (3.81) | 25 993 (2.52) |
| Unknown area | 71 994 (99.40) | 434 (0.60) | 72 428 (7.01) |
| Type of health insurance regime |  |  |  |
| Contributory | 646 670 (97.97) | 13 415 (2.03) | 660 085 (63.89) |
| Subsidized | 146 290 (92.86) | 11 250 (7.14) | 157 540 (15.25) |
| Special | 16 306 (97.24) | 463 (2.76) | 16 769 (1.62) |
| Exception | 47 490 (97.44) | 1247 (2.56) | 48 737 (4.72) |
| Uninsured | 18 447 (97.49) | 474 (2.51) | 18 921 (1.83) |
| Unknown or pending insurance | 10 115 (97.41) | 269 (2.59) | 10 384 (1.01) |
| Non-registered insurance | 117 335 (91.15) | 3447 (2.85) | 120 782 (11.69) |
| Socioeconomic strata |  |  |  |
| 1 Very low | 183 857 (95.51) | 8 644 (4.49) | 192 501 (18.63) |
| 2 Low | 366 446 (96.80) | 12 113 (3.20) | 378 559 (36.64) |
| 3 Middle-low | 200 213 (97.43) | 5 276 (2.57) | 205 489 (19.89) |
| 4 Middle | 35 339 (97.47) | 918 (2.53) | 36 257 (3.51) |
| 5 Middle-high | 11 857 (97.61) | 290 (2.39) | 12 147 (1.18) |
| 6 High | 6126 (96.73) | 207 (3.27) | 6333 (0.61) |
| Non-registered strata | 198 815 (98.46) | 3117 (1.54) | 201 932 (19.54) |

Figure 1 shows the survival functions for each predictor in the model obtained from the multiple Cox Regression without time-dependent factors (See Table S4 in Supplementary material for model details). As the assumption of proportional hazards did not hold for all predictors but sex (see Figure S2-S12 in Supplementary material), we fit the multi-predictor Cox Regression for time dependent variables including the same predictors. This model was statistically significant (p < 0.001). Table 2 presents the results of our final multi-predictor time-dependent Cox regression model (See Tables S5-S6 in supplementary materials for details). The instantaneous risk of dying for COVID-19 among confirmed cases is 59% higher in males compared to females, 27% higher in indigenous people compared to whites/mestizos, and 97% higher in people with subsidized health insurance regime compared to contributory. There was evidence of a dose-response pattern by life-course age groups and SES levels. The risk of dying for COVID-19 among confirmed cases for people over 60 years is extremely higher than the risk for infants. The instantaneous risk of death for people with confirmed diagnosis of COVID-19 living in the very low SES increases by 73% compared to the risk of people living in the high SES (HR=1.73 95% CI: 1.48-2.04). In contrast, living in a sparse rural area decreased the risk of mortality for COVID-19 (HR=0.83 95% CI: 0.76-0.91). Interactions terms between the time to the death and all the variables in the model were statistically significant (p<0.001).

**Table 2.** Risks of death for COVID-19 by some socioeconomic conditions in Colombia up to and including October 26^th^, 2020

| **Socioeconomic condition** | **HR (95% CI)** | **P value** |
| --- | --- | --- |
| Sex (female as reference) | 1.59 (1.53 – 1.65) | <0.001 |
| Age groups (Infants as reference) |  |  |
| Children and school-age | 0.42 (0.22-0.81) | 0.009 |
| Adolescents | 1.16 (0.81-1.67) | 0.414 |
| Young adults | 5.67 (4.00-8.04) | <0.001 |
| Adults | 33.70 (23.59-48.14) | <0.001 |
| Seniors | 214.31 (148.64-309.01) | <0.001 |
| Ethnicity (White/mestizo as reference) |  |  |
| Indigenous | 1.27 (1.13-1.43) | <0.001 |
| Gipsy-Roman | 1.56 (0.39-6.25) | 0.530 |
| Raizal | 0.00 (0.00-3.44) | 0.913 |
| African-Colombian | 1.01 (0.96-1.08) | 0.613 |
| Area of residence (urban as reference) |  |  |
| Semirural (village) | 0.88 (0.82-0.93) | <0.001 |
| Sparse rural | 0.83 (0.76-0.91) | <0.001 |
| Unknown area | 0.14 (0.12-0.16) | <0.001 |
| Type of health insurance regime<br>(contributory regime as reference) |  |  |
| Subsidized | 1.97 (1.89-2.04) | <0.001 |
| Special | 1.29 (1.17-1.41) | <0.001 |
| Exception | 1.37 (1.29-1.45) | <0.001 |
| Uninsured | 1.34 (1.21-1.48) | <0.001 |
| Unknown or pending insurance | 1.22 (1.07-1.39) | 0.002 |
| Non-registered insurance | 2.57 (2.41-2.73) | <0.001 |
| Socioeconomic strata<br>(high SES as reference) |  |  |
| Very low | 1.73 (1.48-2.04) | <0.001 |
| Low | 1.61 (1.38-1.87) | <0.001 |
| Middle-low | 1.34 (1.16-1.56) | <0.001 |
| Middle | 1.16 (0.99-1.36) | 0.059 |
| Middle-high | 0.94 (0.79-1.13) | 0.531 |
| Unknown | 1.54 (1.30-1.83) | <0.001 |

HR: Hazard Ratio; CI: Confidence Interval

**Figure 1.**
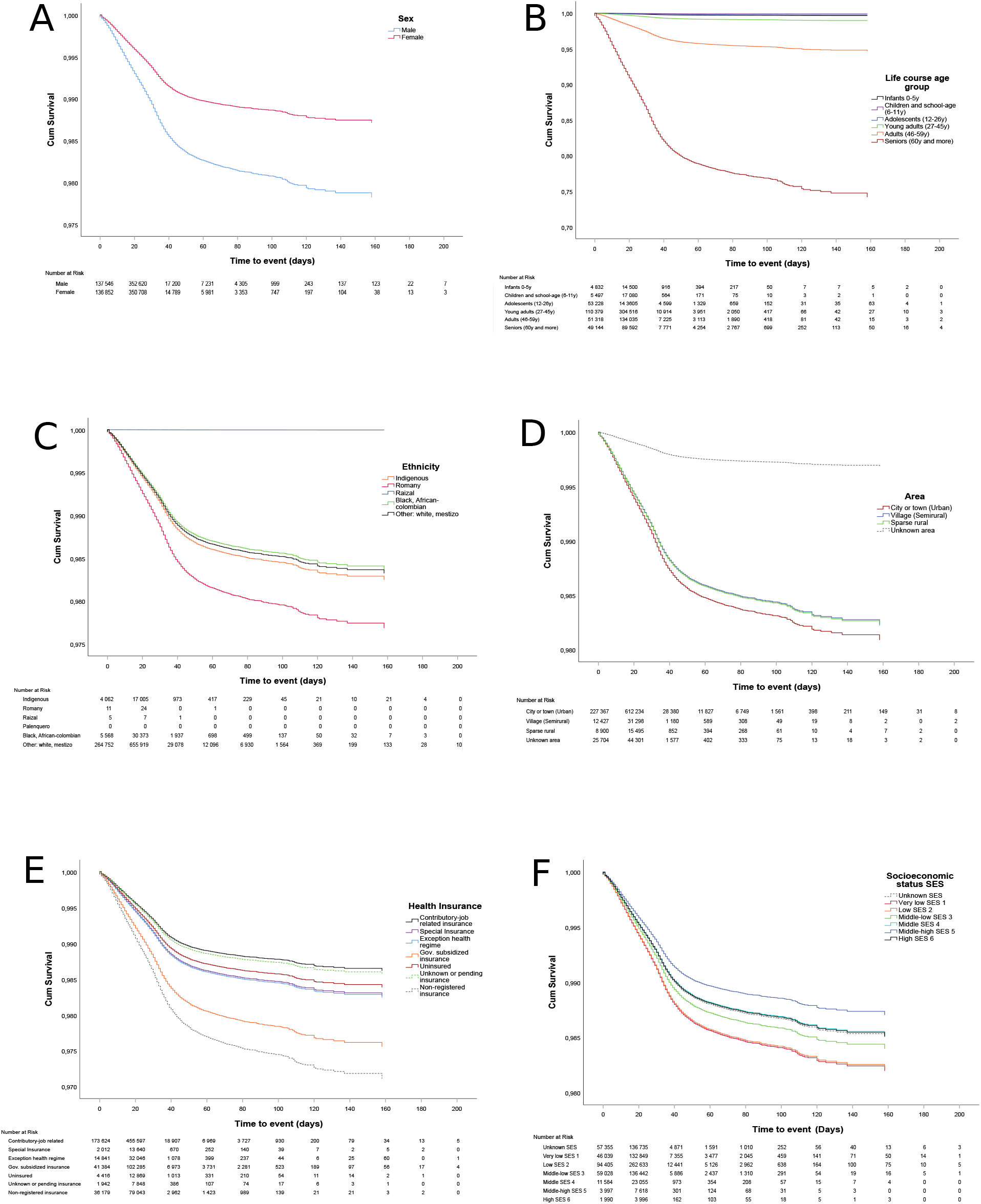
Survival curves for COVID-19 by socioeconomic conditions. A). Sex; B) Age groups; C) Ethnicity; D) Area of residence; E) Health insurance regime; F) Socioeconomic status

## DISCUSSION

Our results provide evidence of socioeconomic and demographic inequalities in COVID-19 mortality in Colombia. In addition to the well documented differential risk of mortality related to older age groups and male sex, this study provides evidence of socioeconomic and ethnicity inequalities in COVID-19 mortality. We identified higher mortality risks for indigenous people, people in the subsidized health regime, and those living in areas classified as very low and low SES. The risks of mortality for age groups and SES levels followed a consistent dose-response pattern.

Our findings of association between COVID-19 mortality and older age (60 year or more) and male sex are consistent with previous reports.[12] The most plausible explanation for this finding is the age-related response to sepsis in older adults with decline in the immune cell function, reduced humoral immune function, and uncontrolled production of inflammatory cytokines.[13] Our study also found an increased risk of death in men which is consistent with previous results. [14] Sex differences in COVID-19 mortality are probably explained by the increased expression in men of the angiotensin converting enzyme-2 (ACE-2), a key factor involved in the pathogenesis of COVID-19.[15]

Ethnicity disparities have been also reported in a variety of contexts. African American and Hispanic in the United States (US) are more vulnerable to COVID-19 mortality than other ethnic groups.[7] In Brazil, after age, Pardo ethnicity was the second most important risk factor for death and probable explanations are differential access to health care or susceptibility to COVID-19 infection.[16] Our study found an increased risk of COVID-19 mortality among indigenous people. Leticia, the capital of the Amazonas department with a live frontier with Brazil, holds the highest COVID-19 mortality rate across departments in Colombia. It is estimated that at least 163 indigenous communities have been infected for the SARS-CoV-2 in Latin America. Poor living and sanitary conditions combined with the burden of previous infectious diseases and malnutrition impose a higher risk to the health of individuals and entire communities.[17]

There is evidence of historical socioeconomic inequalities in previous pandemics. During the 1918 Spanish influenza pandemic there were reports that showed that mortality rates in some countries of South America was 20 times higher compared to countries in Europe.[18] The case fatality rates across countries early in the current COVID-19 pandemic showed negative correlation with countries’ Gross Domestic Product (GDP) and Human Development Index. [19,20] In the US, the COVID-19 pandemic is accelerating the health inequities by disproportionately affecting people from the most disadvantaged groups such as immigrants, people with disabilities and people in prisons and jails.[21] In Brazil, income and education inequalities were positively associated with COVID-19 incidence and mortality rates.[22,23] Our results showed that living in areas of very low or low SES is associated with higher COVID-19 mortality risk with a consistent dose-response effect pattern. These results are consistent with a previous report of inequalities in mortality by SES levels among COVID-19 confirmed cases in Bogotá.[24] These findings are also consistent with results of a nation- wide ecologic study that showed increased risk of COVID-19 mortality associated with the municipalities’ multidimensional poverty index. [25] Colombia has one of the largest income gaps in Latin America and income inequalities within the country differ widely by geographic region in relation to land property, work market, and the effect of violence and armed conflict.[26] These baseline socioeconomic inequalities are translated into higher risk of exposure to and severity of COVID-19 affecting disproportionately to people in lower socioeconomic conditions in Colombia.

Social disruption stress producing pro-inflammatory gene expression has been described as a potential pathological mechanism to explain higher adverse health outcomes in populations with disadvantaged socioeconomic conditions.[27] However, the most possible explanation for the inequalities in COVID-19 mortality are the historical inequalities in terms of living and working conditions, and the unequal access to health care services. Inequalities in working conditions might explain an important part of the inequalities of COVID-19 infection and mortality. In our study, people in the subsidized health insurance regime represent people with unstable or informal work, or unemployed people who need subsidy from the government to get access to health services. Thus, the higher mortality risk observed in this group compared to the contributory health regime might be representing the social inequality related to working conditions in Colombia. People in the more disadvantaged working groups have lower-paid work and are more likely to work in key basic services (food, cleaning, delivery or public services) that require them to work in person and commute across the cities.[28] In contrast, people with higher-paid work are more likely to work from home with lower exposure to COVID-19 infection.[29]

Despite having an almost universal health insurance coverage, the Colombian health system is characterized by a strong fragmentation in the provision of health care services, an incipient primary health care, and differences in quality of health care services across regimes.[30] Therefore, differences between contributory and subsidized groups might be explained not only by underlying working conditions but also for chronic inequalities in access to high quality health care services. Limited health care services are provided in semi-rural and sparse rural areas. Our findings, however, found a potential protector effect for COVID-19 mortality for people living in those areas compared to people living in urban areas. The direction of this association might be explained by the SARS-CoV-2 transmission dynamics that favors contagion among close contacts in crowded places which is more common in urban areas.

The COVID-19 pandemic is occurring in presence of a non-communicable diseases (NCDs) epidemic and within a context of historical inequalities in the social determinants of health, which is recognized as a syndemic.[29] There are complex connections among NCDs, COVID-19 transmission dynamics and living conditions that shape disparities with higher adverse effects for disadvantaged people. People from minority ethnic groups, people living in areas with higher socioeconomic deprivation, generally have a greater number of or more severe or uncontrolled coexisting NCDs.[31] These inequalities in chronic conditions are deepened by the way people live and work which make them also more exposed to COVID-19 infection and mortality. Therefore, there is a need to measure, analyze and report demographic and socioeconomic inequities for identifying groups at higher risk for COVID-19 mortality in order to guide tailored public health interventions in countries.[8]

Our study provides strong evidence of socioeconomic inequalities in COVID-19 mortality in Colombia by using data from a nation-wide cohort of confirmed cases during the first eight months of the epidemic. However, conclusions should be carefully interpreted considering the limitations of the study. This study relies on data reported to SIVIGILA and it is possible that despite its national coverage, some degree of underreporting might be present. The probability of underreporting is higher in the sparse rural areas where most disadvantaged people live and therefore underreporting, if present, would have an attenuating effect of the effect measures. Colombia does not conduct COVID-19 mass testing and underreporting of cases and deaths might be influencing the confirmed cases captured by SIVIGILA and our results. Finally, our results are not controlled for the presence of chronic morbidities in confirmed cases so the effect of specific chronic diseases on COVID-19 mortality was not estimated and socioeconomic variables are not controlled for them.

In conclusion, our study provides evidence of demographic and socioeconomic inequalities in COVID-19 mortality in terms of age groups, sex, ethnicity, type of health insurance regime, and socioeconomic strata. Confirmed COVID-19 cases who are male, over 60 years old, indigenous, holding a government subsidized health insurance, and those living in areas classified in the lower socioeconomic strata have a higher risk of dying faster from COVID-19. Public health interventions for prevention and early detection of COVID-19 cases should be prioritized for more vulnerable groups according to the unequal mortality risks.

## Data Availability

Data used for the current study are publicly available as open data on the government website
 

https://www.datos.gov.co/en/Salud-y-Protecci-n-Social/Casos-positivos-de-COVID-19-en-Colombia/gt2j-8ykr

## Contributor’s statement

MPC: methodology design, verification of the underlying data, data analysis, data interpretation, writing-original draft

LAR-V: literature research, data analysis, data interpretation, writing-original draft

MLR-B: verification of the underlying data, data analysis, data interpretation, writing – review and editing

CA-M: data interpretation, writing – review and editing

JAF-N: conceptualization, methodology design, data analysis, data interpretation, writing – review and editing

## Data Availability Statement

Data used for the current study are publicly available as open data on the government website (https://www.datos.gov.co/en/Salud-y-Protecci-n-Social/Casos-positivos-de-COVID-19-en-Colombia/gt2j-8ykr)

## Supplemental Figures and Tables

**Figure S1.**
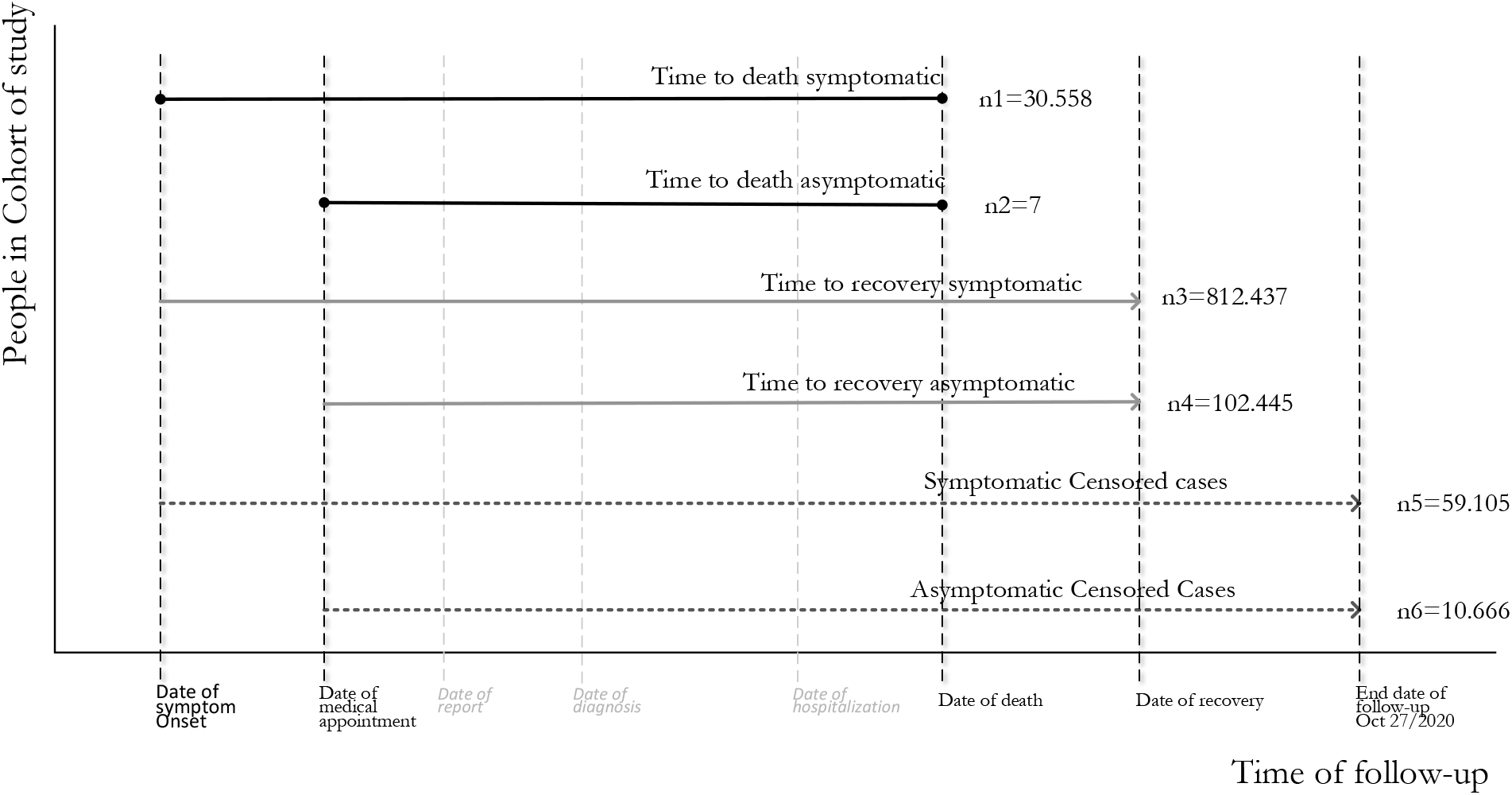
Approaches to compute days of time to event in the cohort of COVID-19 confirmed cases, Colombia.

**Table S1.** Descriptive statistics and distributions of the six times to death, to recovery and censored cases, each for symptomatic and asymptomatic cases.

|  | Time to event | Time to death symptomatic | Time to death, asymptomatic | Time to recovery, symptomatic | Time to recovery, asymptomatic | Symptomatic censored cases | Asymptomatic censored cases |
| --- | --- | --- | --- | --- | --- | --- | --- |
| N | 1 033 218 | 30 558 | 7 | 812 437 | 120 445 | 59 105 | 10 666 |
| Mean | 25.56 | 15.97 | 16.43 | 25.92 | 25.64 | 28.00 | 11.56 |
| Std. Error of Mean | .01 | .07 | 4.61 | .01 | .02 | .12 | .12 |
| Median | 24.00 | 14.00 | 11.00 | 25.00 | 25.00 | 14.00 | 10.00 |
| Mode | 21 | 10 | 8 | 21 | 25 | 14 | 13 |
| Std. Deviation | 11.81 | 11.88 | 12.20 | 9.37 | 8.53 | 29.89 | 12.81 |
| Variance | 139.62 | 141.04 | 148.95 | 87.74 | 72.69 | 893.26 | 164.19 |
| Range | 216 | 158 | 34 | 211 | 196 | 215 | 213 |
| Minimum | 0 | 0 | 8 | 0 | 2 | 1 | 2 |
| Maximum | 216 | 158 | 42 | 211 | 198 | 216 | 215 |
| Sum | 26 411.767 | 488 051 | 115 | 21 056 824 | 3 088 341 | 1 655 125 | 123.311 |
| Percentiles |  |  |  |  |  |  |  |
| 25 | 20 | 8 | 8 | 21 | 20 | 10 | 7 |
| 50 | 24 | 14 | 11 | 25 | 25 | 14 | 10 |
| 75 | 29 | 22 | 20 | 29 | 29 | 35 | 12 |
| IQR | 9 | 14 | 12 | 8 | 9 | 15 | 5 |

**Table S2.** Frequencies of predictors’categories by type of time-to-event outcome^1^

| Predictors / categories | Time to event outcomes |  |  |  |  |  |
| --- | --- | --- | --- | --- | --- | --- |
|  | 1 | 2 | 3 | 4 | 5 | 6 |
| **Sex** |  |  |  |  |  |  |
| Male | 19 608 | 5 | 405 391 | 59 694 | 30 547 | 5 188 |
| Female | 10 950 | 2 | 407 046 | 60 751 | 28 558 | 5 478 |
| **Socioeconomic status SES** |  |  |  |  |  |  |
| Unknown SES | 3 115 | 2 | 148 867 | 33 125 | 11 229 | 5 594 |
| Very low SES 1 | 8 644 | 0 | 158 065 | 13 029 | 12 236 | 527 |
| Low SES 2 | 12 110 | 3 | 303 406 | 40 542 | 20 541 | 1 957 |
| Middle-low SES 3 | 5 275 | 1 | 160 508 | 26 072 | 11 741 | 1 892 |
| Middle SES 4 | 917 | 1 | 27 691 | 5 004 | 2 200 | 444 |
| Middle-high SES 5 | 290 | 0 | 9 289 | 1 672 | 732 | 164 |
| High SES 6 | 207 | 0 | 4 611 | 1 001 | 426 | 88 |
| **Life course age group** |  |  |  |  |  |  |
| Infants 0-5y | 38 | 0 | 16 068 | 3 096 | 1 504 | 224 |
| Children and school-age (6-11y) | 12 | 0 | 18 147 | 3 989 | 985 | 270 |
| Adolescents (12-26y) | 230 | 1 | 164 869 | 26 568 | 9 821 | 2 217 |
| Young adults (27-45y) | 2 000 | 0 | 355 500 | 49 830 | 20 775 | 4 270 |
| Adults (46-59y) | 5 000 | 0 | 156 162 | 2 2847 | 12 094 | 2 039 |
| Seniors (60y and more) | 23 278 | 6 | 101 691 | 14 115 | 13 926 | 1 646 |
| **Ethnicity** |  |  |  |  |  |  |
| Indigenous | 776 | 0 | 19 502 | 1 851 | 621 | 37 |
| Romany | 2 | 0 | 30 | 3 | 1 | 0 |
| Raizal | 0 | 0 | 5 | 8 | 0 | 0 |
| Palenquero | 0 | 0 | 0 | 0 | 0 | 0 |
| Black, African-colombian | 1 421 | 0 | 34 901 | 2 339 | 637 | 6 |
| Other: white, mestizo | 28 359 | 7 | 757999 | 116 244 | 57 846 | 10 623 |
| **Area** |  |  |  |  |  |  |
| City or town (Urban) | 27 837 | 7 | 708 770 | 98 609 | 48 541 | 5 151 |
| Village (Semirural) | 1 296 | 0 | 39 335 | 1 946 | 3226 | 79 |
| Sparse rural | 991 | 0 | 21 322 | 673 | 2972 | 35 |
| Unknown area | 434 | 0 | 43 010 | 19 217 | 4366 | 5 401 |
| **Health Insurance** |  |  |  |  |  |  |
| Contributory-job related insurance | 13 411 | 4 | 527 518 | 80 960 | 33700 | 4 492 |
| Special Insurance | 463 | 0 | 14 274 | 1 739 | 275 | 18 |
| Exception health regime | 1245 | 2 | 40 636 | 3 533 | 3089 | 232 |
| Gov. subsidized insurance | 11 249 | 1 | 124 099 | 8 988 | 12837 | 366 |
| Uninsured | 474 | 0 | 13 485 | 3 671 | 1214 | 77 |
| Unknown or pending insurance | 269 | 0 | 8 821 | 778 | 505 | 11 |
| Non-registered insurance | 3 447 | 0 | 83 604 | 20 776 | 7485 | 5 470 |
| **TOTAL** | **30 558** | **7** | **812 437** | **120 445** | **59105** | **10 666** |

<sup>1</sup>Time to event outcome 1 refers to time to death of symptomatic cases, and outcome 2 to time to death of asymptomatic ones. Outcome 3 and 4 refer to the frequencies of time to recovery on patients with or without symptoms respectively, and outcome 5 and 6 refer to frequencies of censored cases, also with or without symptoms, respectively.

**Table S3.**
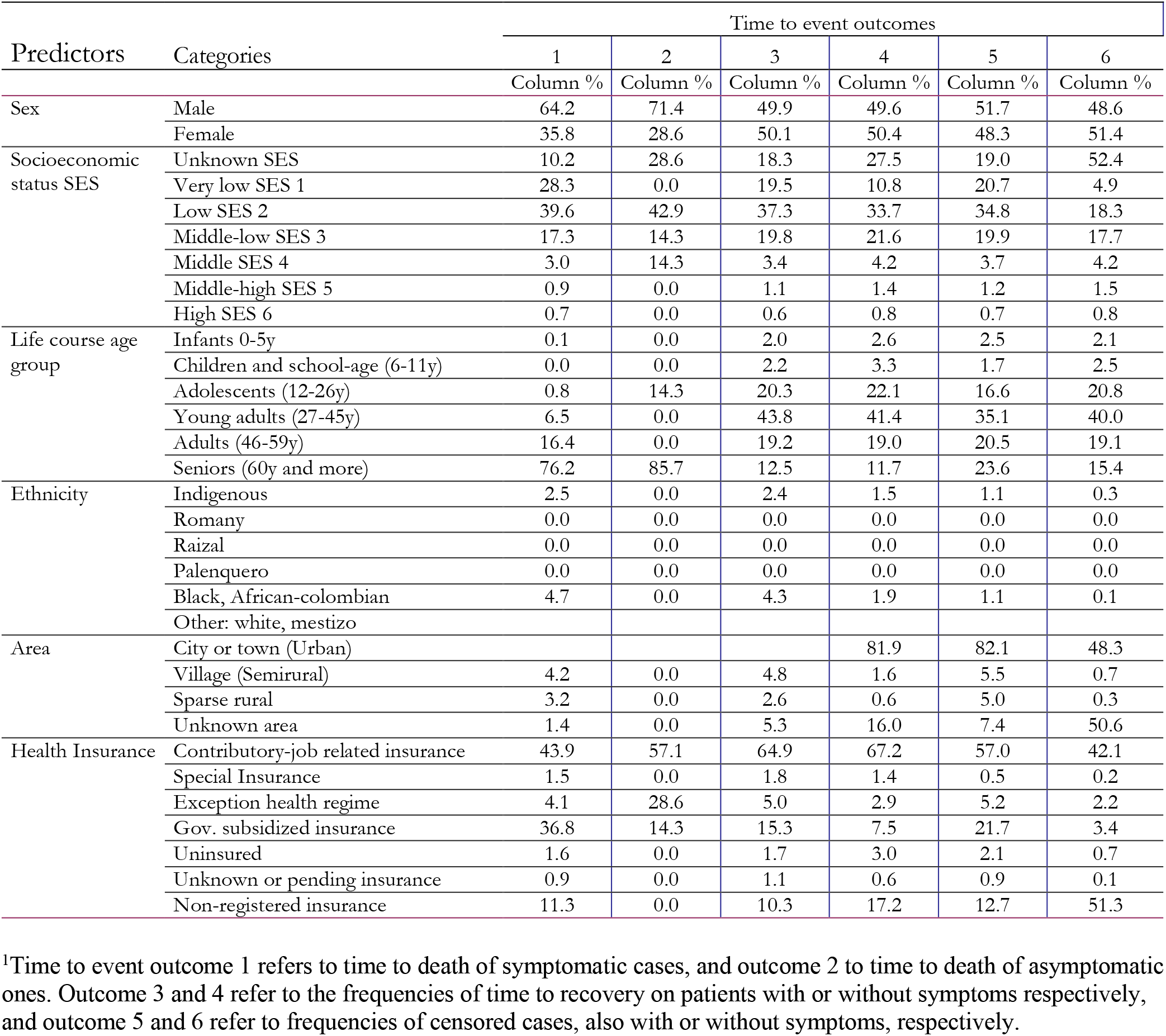
Proportional distribution of predictors’categories by type of time-to-event outcome^1^

**Table S4.** Multi-predictor Cox Regression model without terms to address time-dependent factors

|  | Variables in the Equation |  |  |  |  |  | 95.0% CI for |  |
| --- | --- | --- | --- | --- | --- | --- | --- | --- |
|  | B | SE | Wald | df | Sig. | Exp(B) | Exp(B) |  |
|  |  |  |  |  |  |  | Lower | Upper |
| Sex (Ref. Female) | .53 | .01 | 1 968.10 | 1 | .000 | 1.70 | 1.66 | 1.74 |
| Socioeconomic status SES (Ref. High SES 6) |  |  | 147.55 | 6 | .000 |  |  |  |
| Unknown SES | .01 | .07 | .02 | 1 | .899 | 1.01 | .88 | 1.16 |
| Very low SES 1 | .19 | .07 | 7.43 | 1 | .006 | 1.21 | 1.06 | 1.39 |
| Low SES 2 | .18 | .07 | 6.92 | 1 | .009 | 1.20 | 1.05 | 1.38 |
| Middle-low SES 3 | .07 | .07 | 1.05 | 1 | .305 | 1.08 | .94 | 1.24 |
| Middle SES 4 | -.01 | .08 | .00 | 1 | .946 | .99 | .86 | 1.16 |
| Middle-high SES 5 | -.14 | .09 | 2.38 | 1 | .123 | .87 | .73 | 1.04 |
| Life course age (Infants 0-5y) |  |  | 32 895.07 | 5 | .000 |  |  |  |
| Children and school-age (6-11y) | -1.19 | .33 | 13.02 | 1 | .000 | .30 | .16 | .58 |
| Adolescents (12-26y) | -.35 | .18 | 4.06 | 1 | .044 | .70 | .50 | .99 |
| Young adults (27-45y) | 1.07 | .16 | 42.99 | 1 | .000 | 2.93 | 2.12 | 4.03 |
| Adults (46-59y) | 2.71 | .16 | 276.42 | 1 | .000 | 14.99 | 10.89 | 20.63 |
| Seniors (60y and more) | 4.40 | .16 | 735.60 | 1 | .000 | 81.74 | 59.46 | 112.37 |
| Ethnicity (Ref. Other. white. mestizo) |  |  | 2.85 | 4 | .582 |  |  |  |
| Indigenous | .05 | .04 | 1.51 | 1 | .219 | 1.05 | .97 | 1.12 |
| Romany | .33 | .71 | .21 | 1 | .645 | 1.39 | .35 | 5.54 |
| Raizal | -6.22 | 57.11 | .01 | 1 | .913 | .00 | .00 | 8.09E+45 |
| African-Colombian | -.03 | .03 | 1.01 | 1 | .315 | .97 | .92 | 1.03 |
| Area (ref. City or town (Urban)) |  |  | 1 109.95 | 3 | .000 |  |  |  |
| Village (Semirural) | -.08 | .03 | 7.52 | 1 | .006 | .92 | .87 | .98 |
| Sparse rural | -.07 | .03 | 4.82 | 1 | .028 | .93 | .87 | .99 |
| Unknown area | -1.82 | .05 | 1 104.14 | 1 | .000 | .16 | .15 | .18 |
| Health Insurance (ref. job related insurance) |  |  | 2 274.11 | 6 | .000 |  |  |  |
| Special Insurance | .23 | .05 | 23.06 | 1 | .000 | 1.26 | 1.14 | 1.38 |
| Exception health regime | .24 | .03 | 65.57 | 1 | .000 | 1.27 | 1.20 | 1.35 |
| Gov. subsidized insurance | .57 | .01 | 1 622.47 | 1 | .000 | 1.78 | 1.73 | 1.83 |
| Uninsured | .16 | .05 | 11.07 | 1 | .001 | 1.17 | 1.07 | 1.28 |
| Unknown or pending insurance | .04 | .06 | .34 | 1 | .562 | 1.04 | .92 | 1.17 |
| Non-registered insurance | .74 | .02 | 1 327.97 | 1 | .000 | 2.10 | 2.02 | 2.19 |

### Testing of Assumptions

Additional to overall model statistics, and to systematically explore quantitative criteria of time dependent variables in the model (see interactions between predictors and time in the final Cox Regression with time dependent variables), we used graphical criteria to assess if proportional hazards hold. The following figures present plots between log minus log transformation of survival estimates and time to event, and scatter plots between partial residuals and time to event.

**Figure S2.**
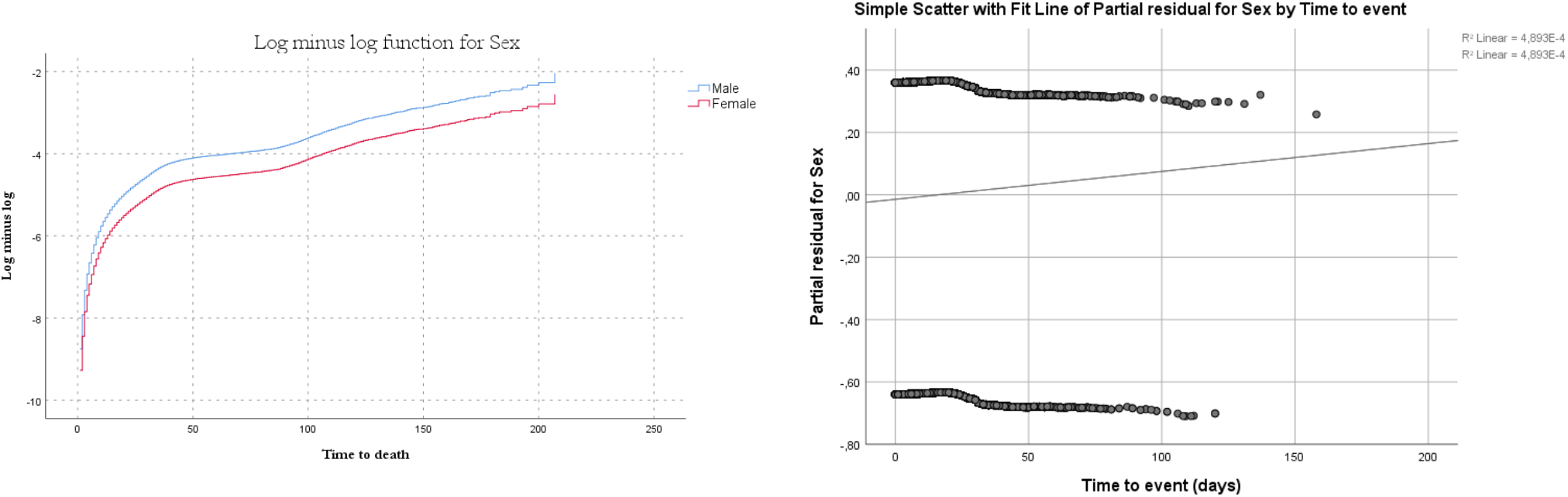
Graphic test of proportional hazard assumption for Sex. Left, Ln(-ln) transformation of survival estimate vs Time to event plot. Right, Partial residual vs time to event plot.

### Sex

Despite the Log minus log plot show almost parallel lines for male an female categories, the partial residual plot shows a positive trend. The interaction between Sex and time to event has a significative p-value close to zero.

### Socioeconomic status

Curves of Ln(-ln) transformation of survival estimates for SES categories do not overlap or cross. However, partial residual plots for all categories but Middle-high SES, have clear trends. Additionally, as interaction between SES and time to event had a p-value close to zero, we conclude SES violates the proportional hazards assumption.

**Figure S3.**
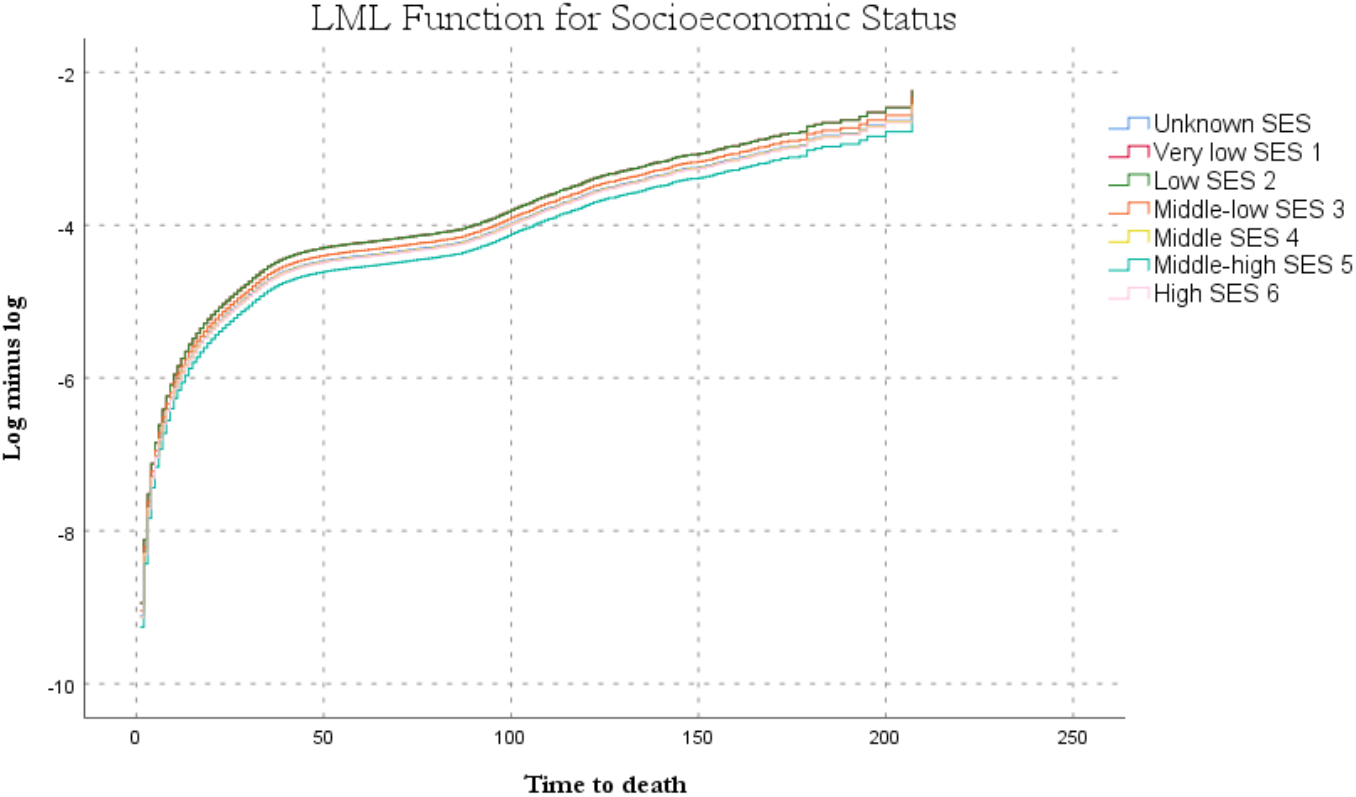
Ln(-ln) transformation of survival estimates for SES categories vs Time to event plot.

**Figure S4.**
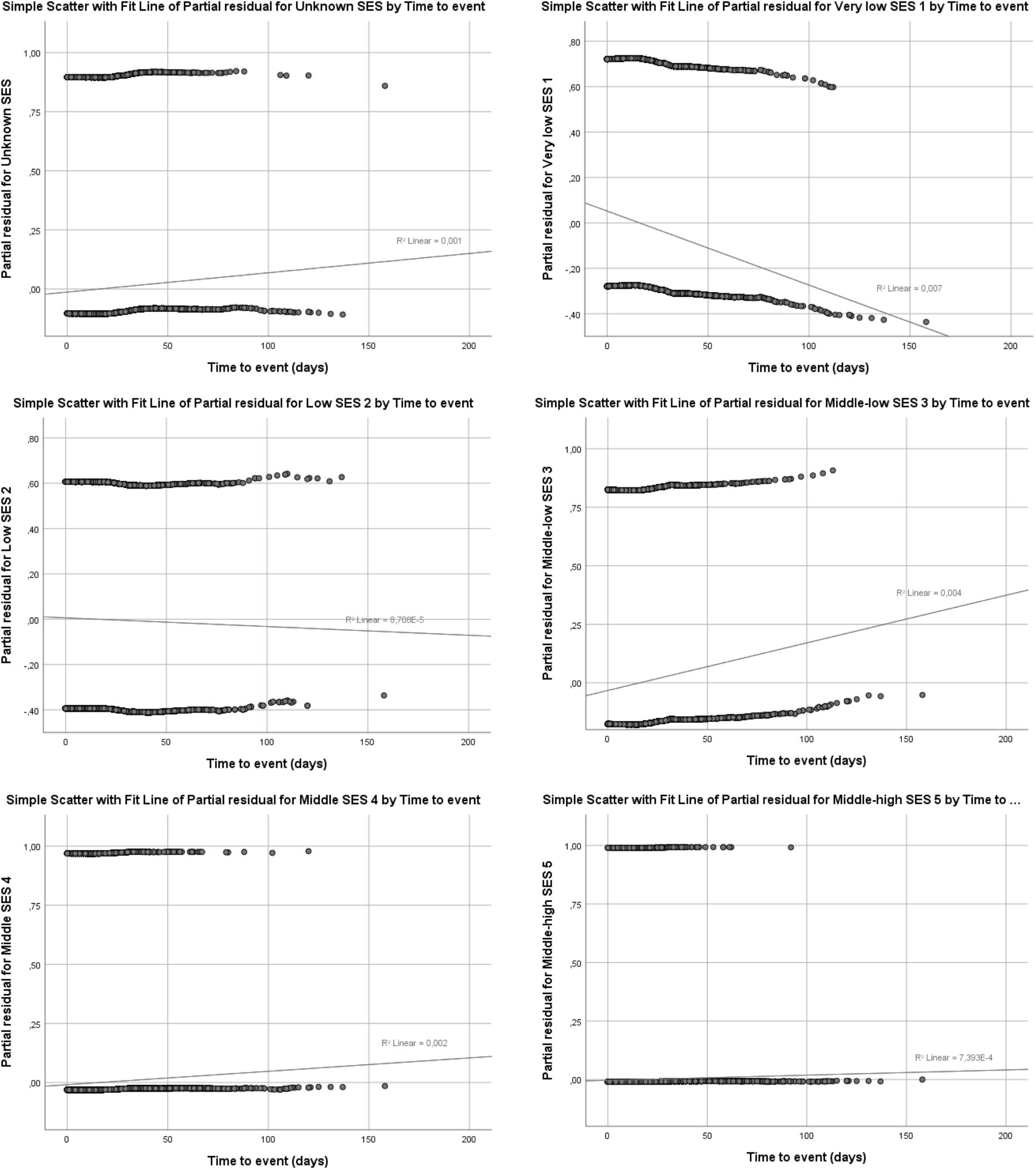
Partial residual vs time to event plots for SES categories.

### Life Course age groups

Similar to the former variables, the log minus log curves for categories in Life course just converge in the origin, with no further crossings. However, partial residual plots have clear trends for categories of young adults, adults and seniors, and for the interaction between Life course age variable and time to event has an overall p-value is close to zero, which confirms proportional hazards assumption do not hold.

**Figure S5.**
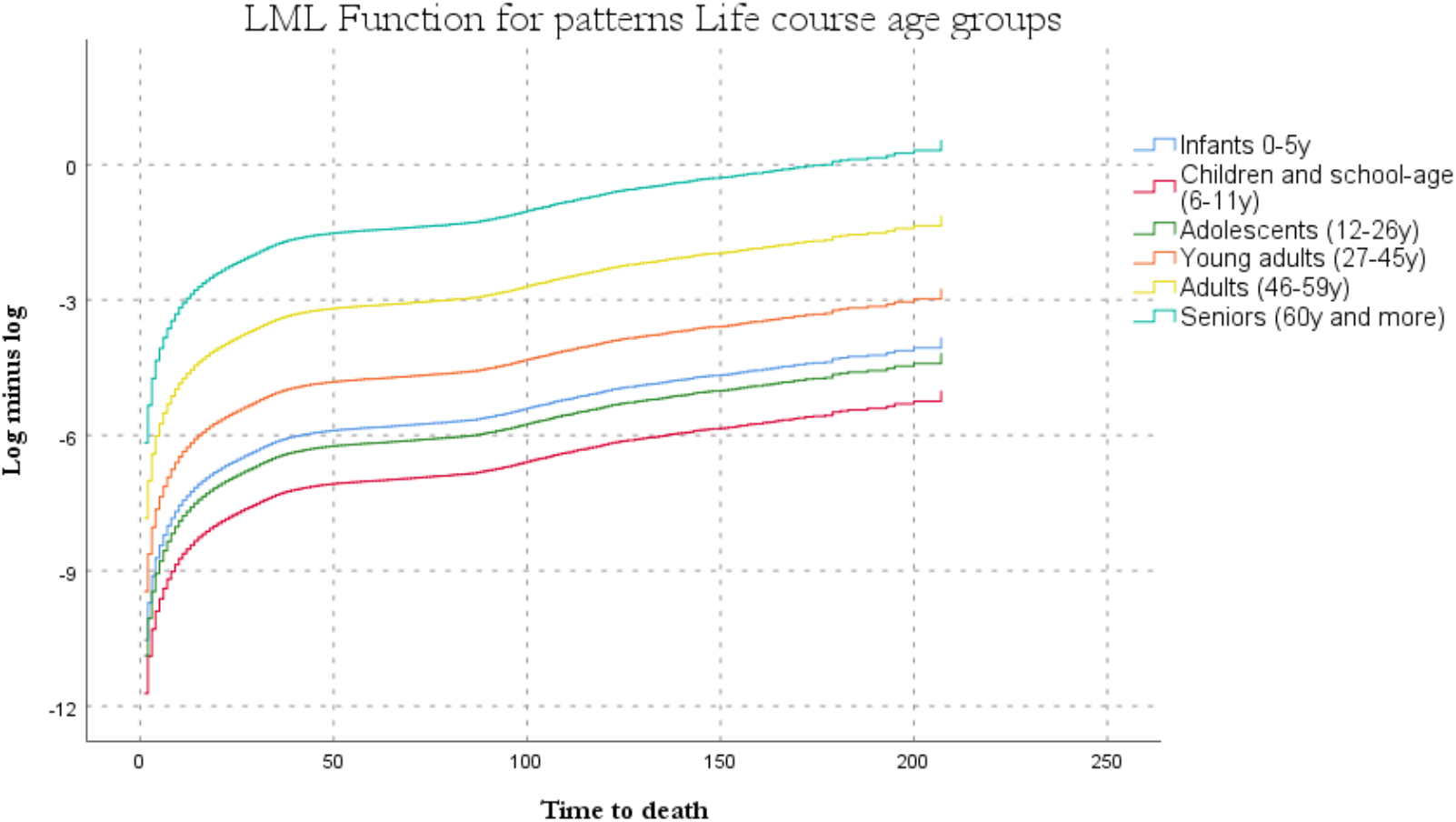
Ln(-ln) transformation of survival estimates for Life course age groups vs Time to event plot.

**Figure S6.**
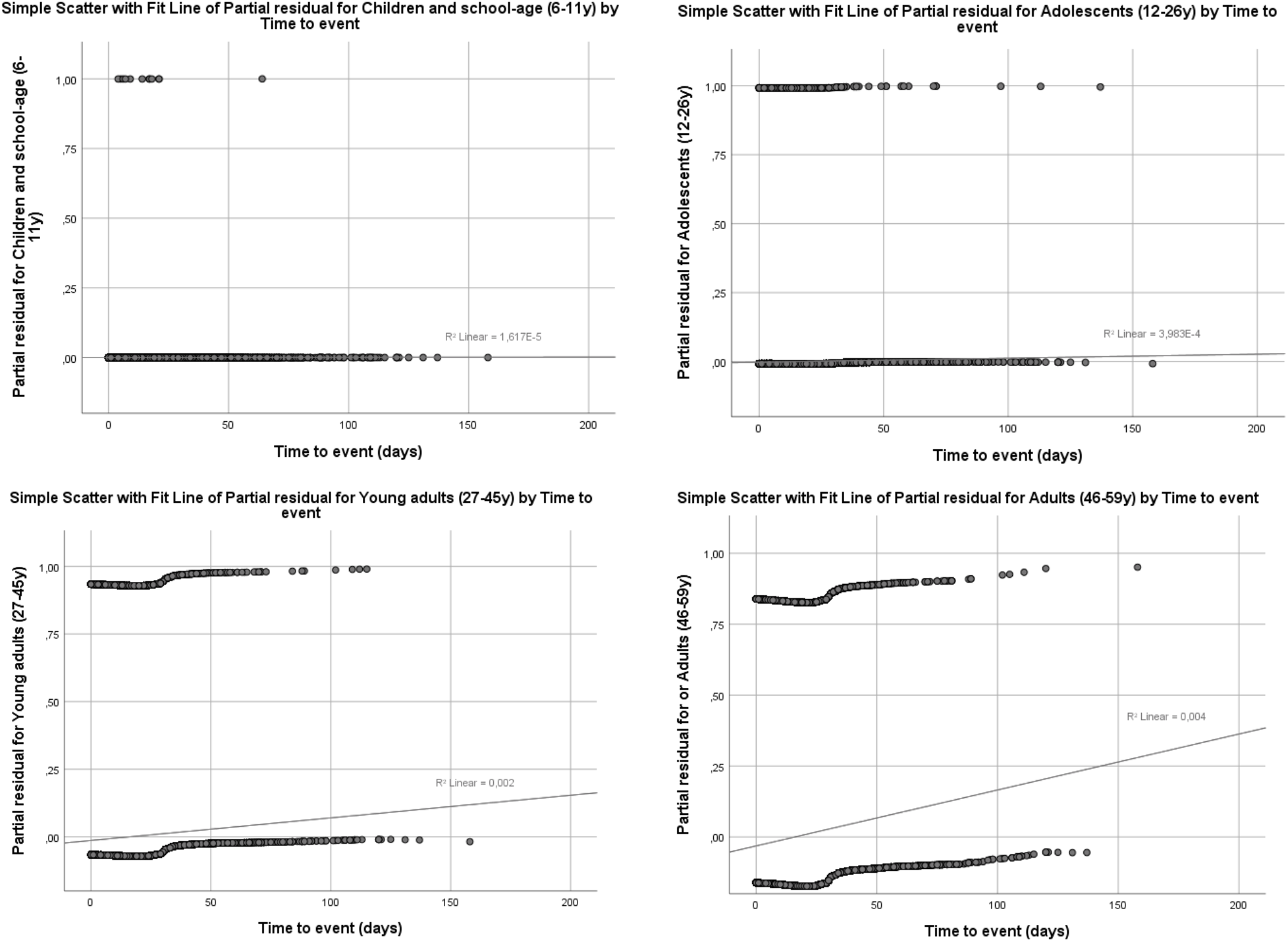
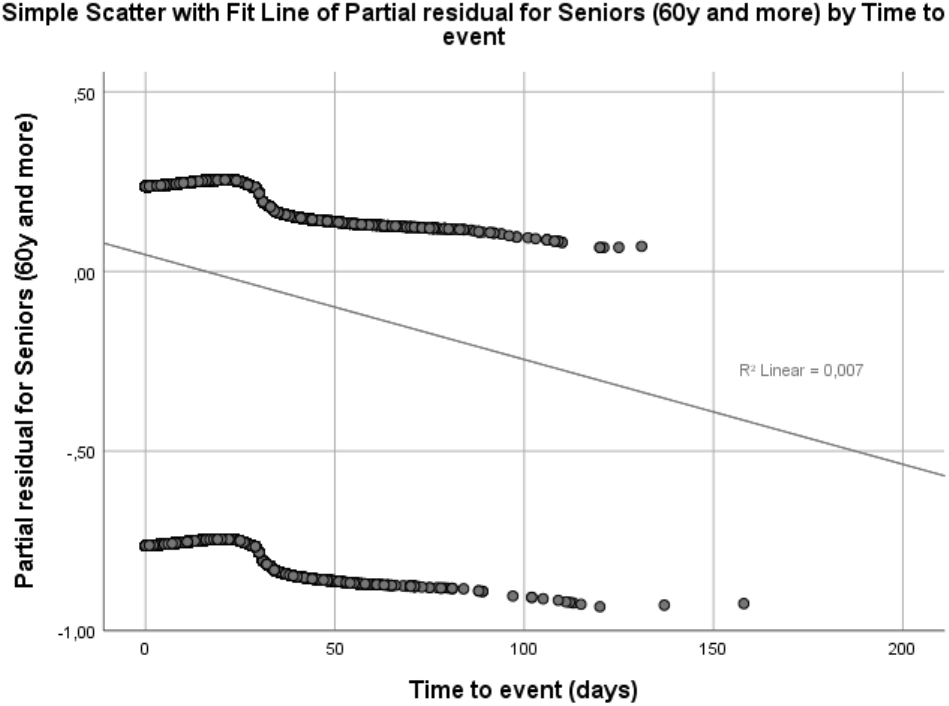
Partial residual vs time to event plots for Life course age groups.

### Ethnicity

The shape of log minus log curves for categories in the ethnicity variable was parallel with no evident crossings. However, partial residual plots for Indigenous, Raizal and Black or African-Colombian groups, had a negative trend, and the p-value o the interaction between ethnicity and time to event close to zero, showed significant deviations from the expected assumption of proportional hazards.

**Figure S7.**
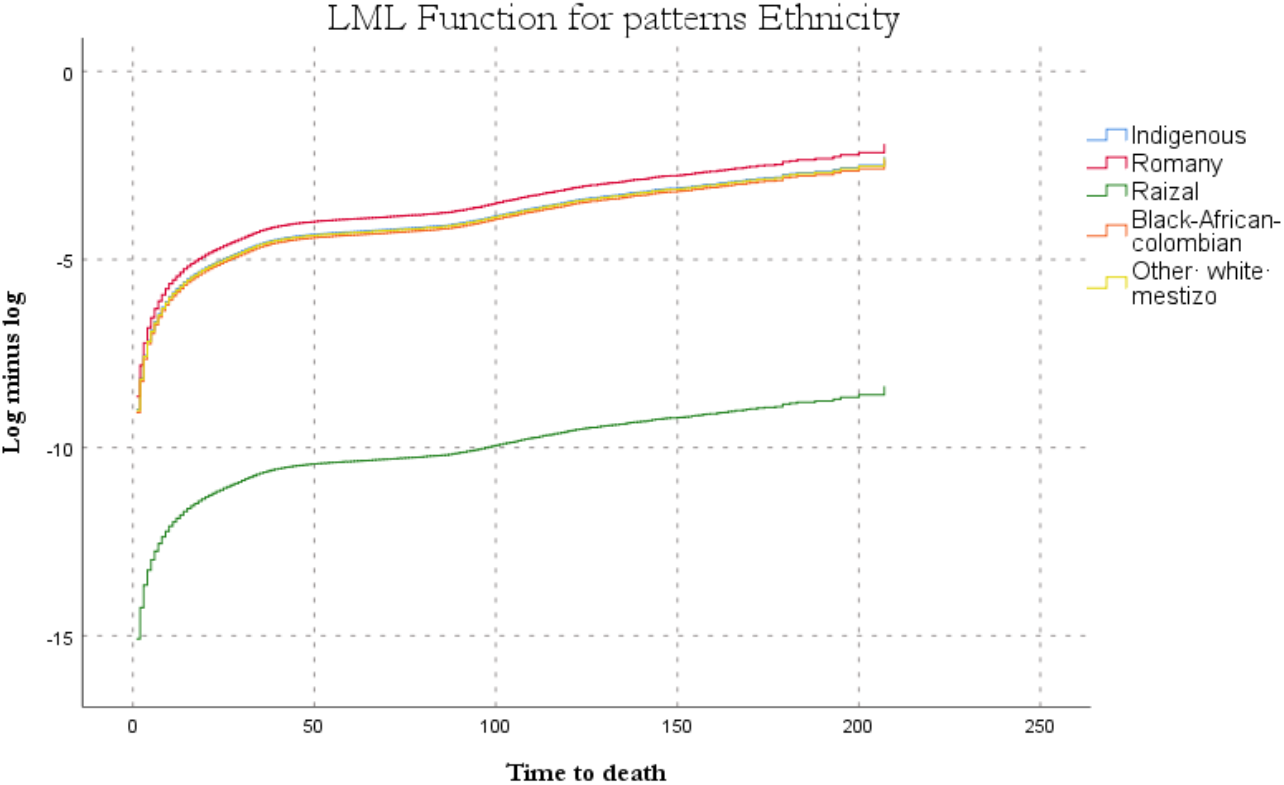
Ln(-ln) transformation of survival estimates for Ethnicity vs Time to event plot.

**Figure S8.**
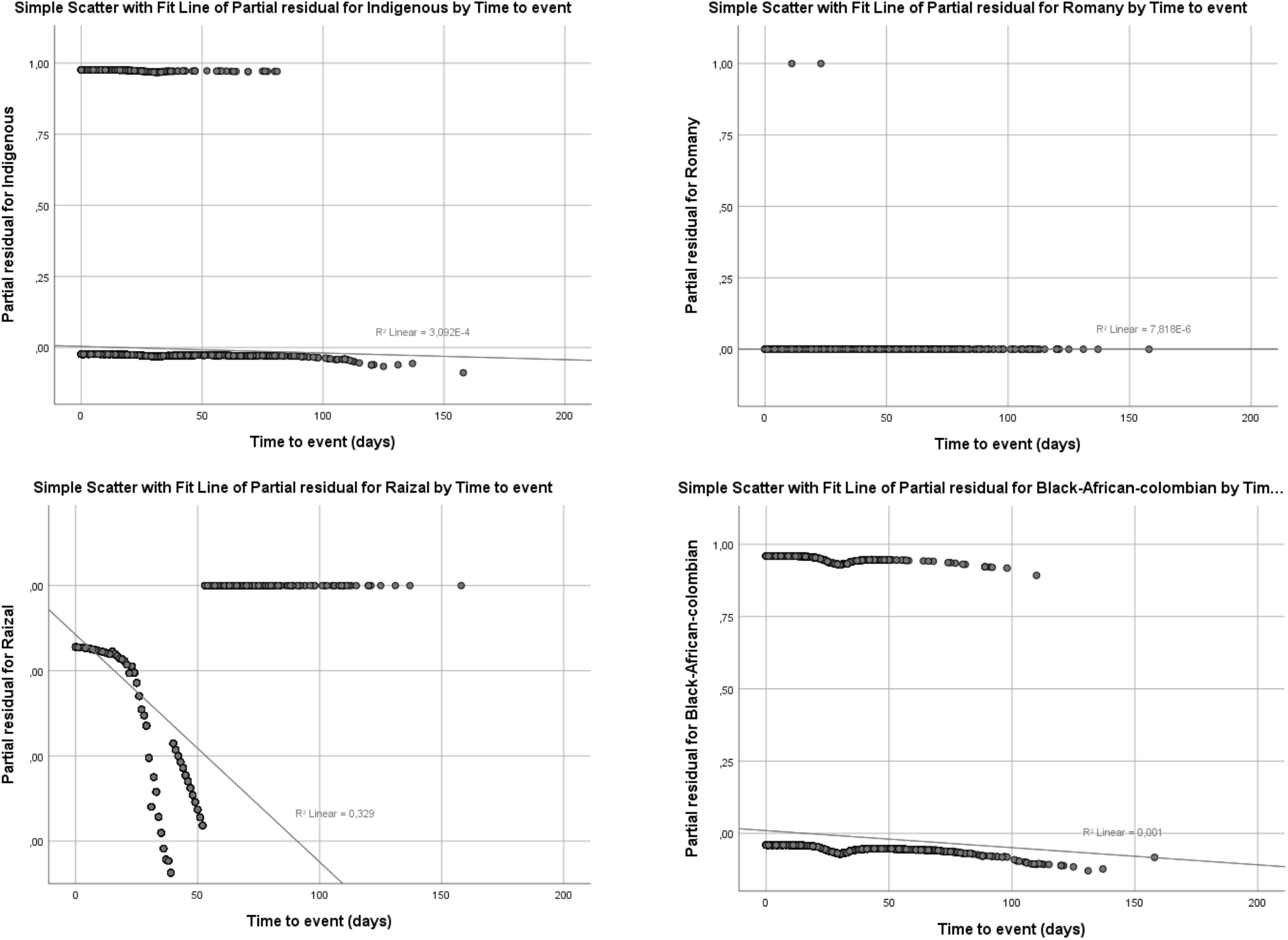
Partial residual vs time to event plots for Ethnicity.

### Area of residence

Almost parallel log minus log curves and almost flat partial residual plots contrast with the close to zero p-value of the interaction term between Area and time to event. By prioritizing the quantitative criterion, we opted to include this interaction in the final model.

**Figure S9.**
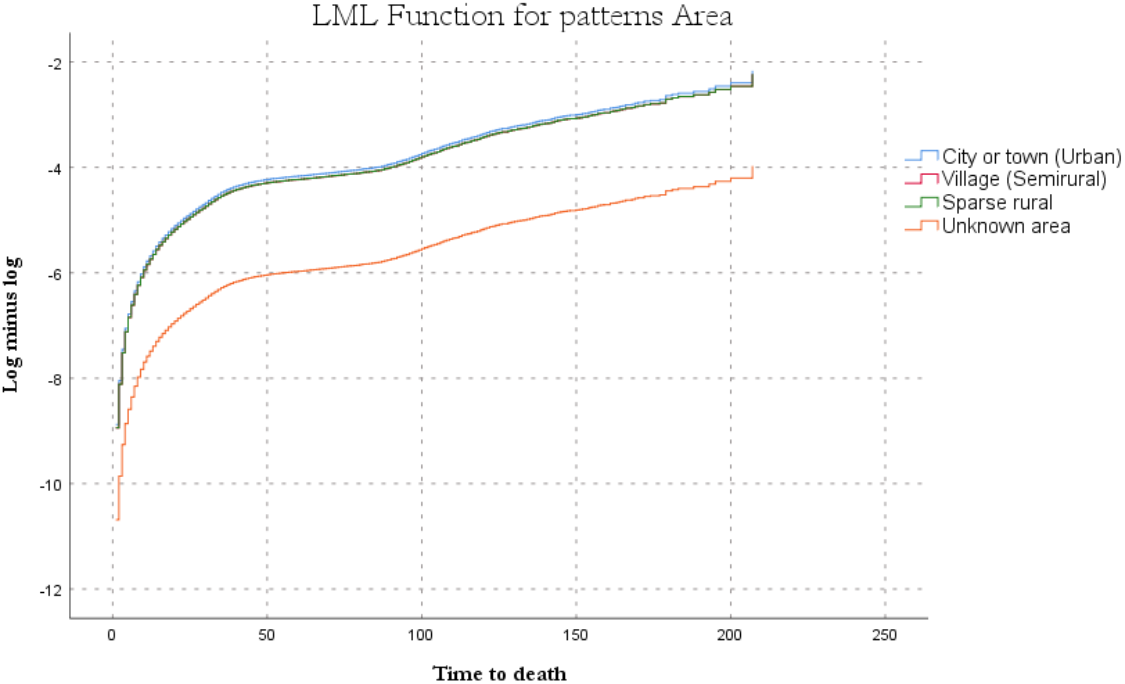
Ln(-ln) transformation of survival estimates for Area vs Time to event plot.

**Figure S10.**
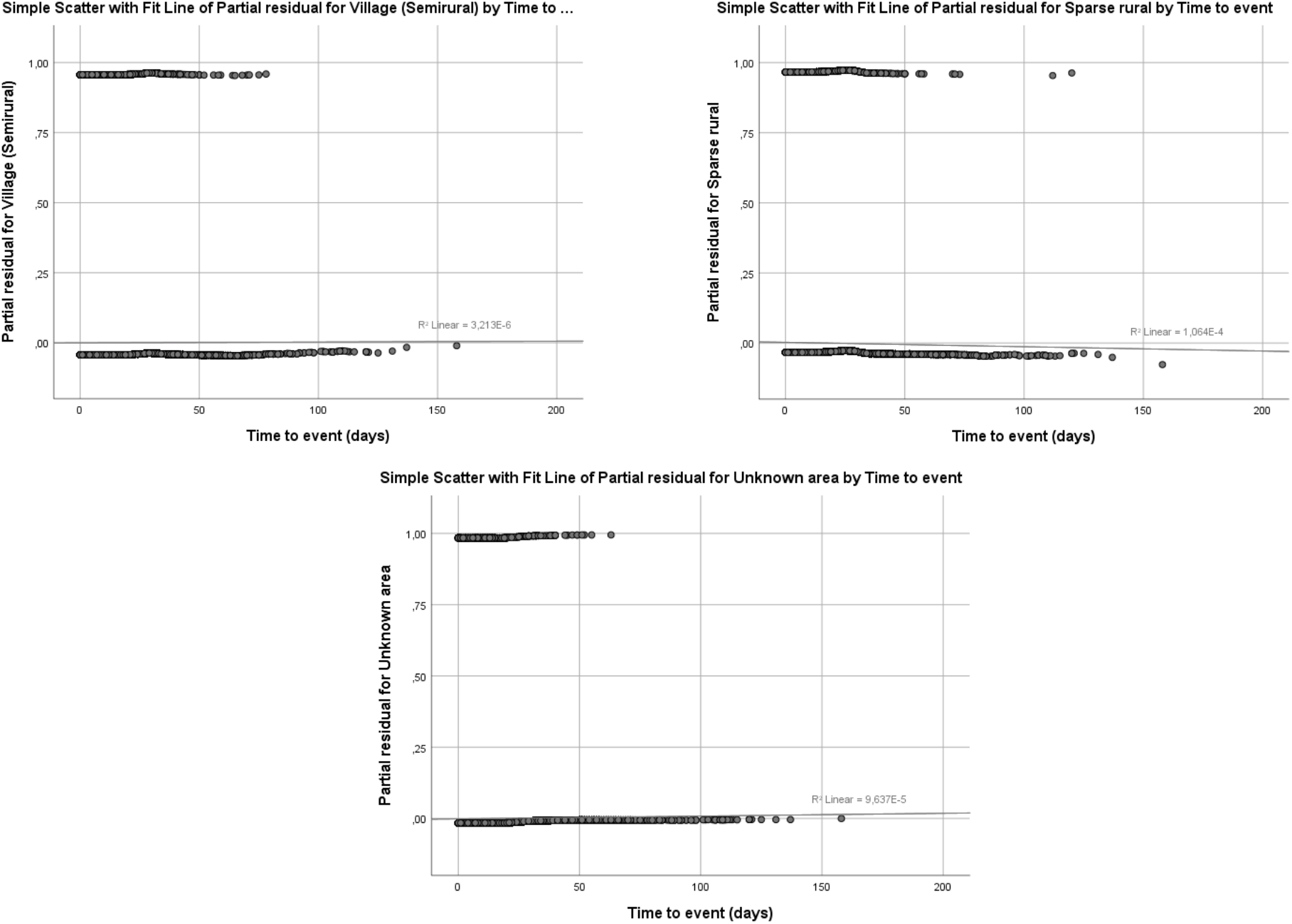
Partial residual vs time to event plots for Area.

### Health insurance

Almost parallel log minus log curves for health insurance categories, which just show convergence at the origin, contrast with marked trends on partial residual plots for Subsidized insurance and the group of people with no registries of the type of health insurance. Exception regime had a slight positive trend and special regime had a slight negative trend. The overall p-value of the interaction between health insurance and time to event was also close to zero.

**Figure S11.**
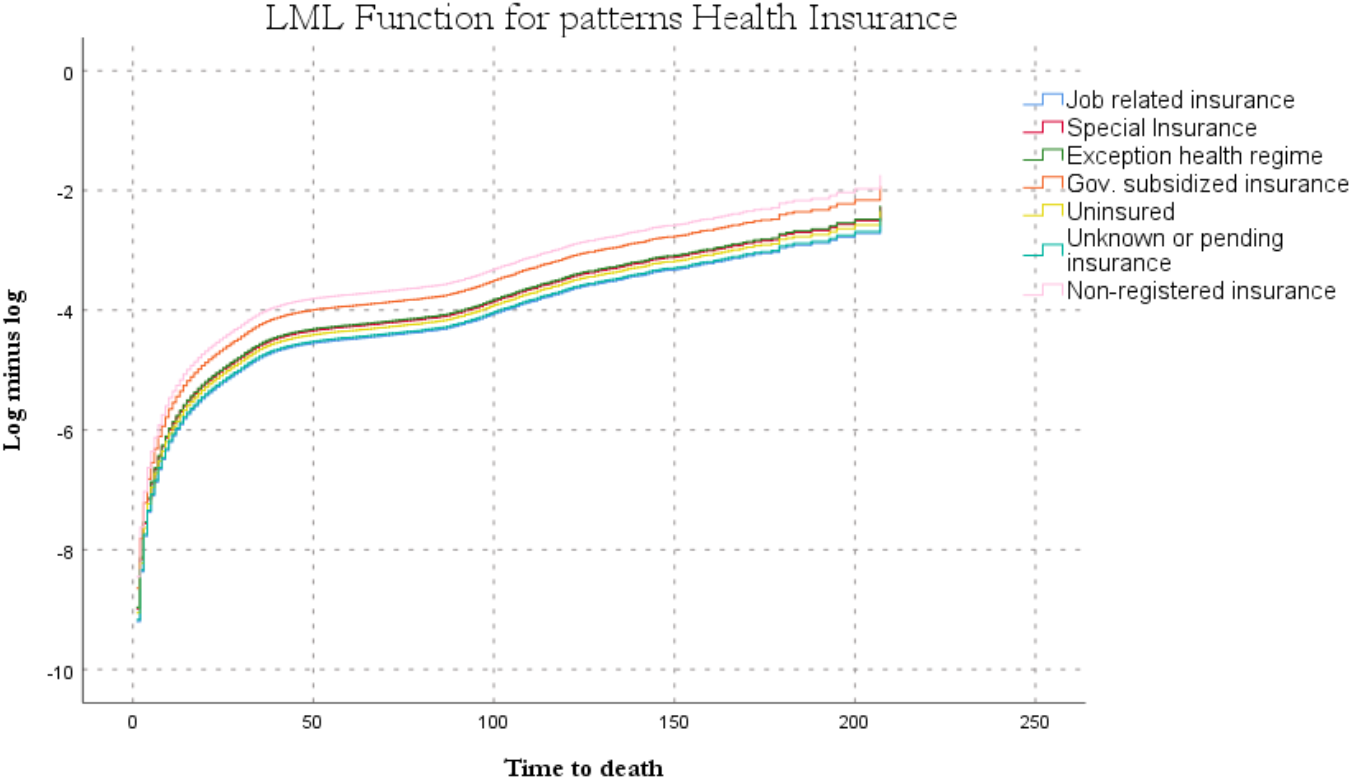
Ln(-ln) transformation of survival estimates for Health insurance vs Time to event plot.

**Figure S12.**
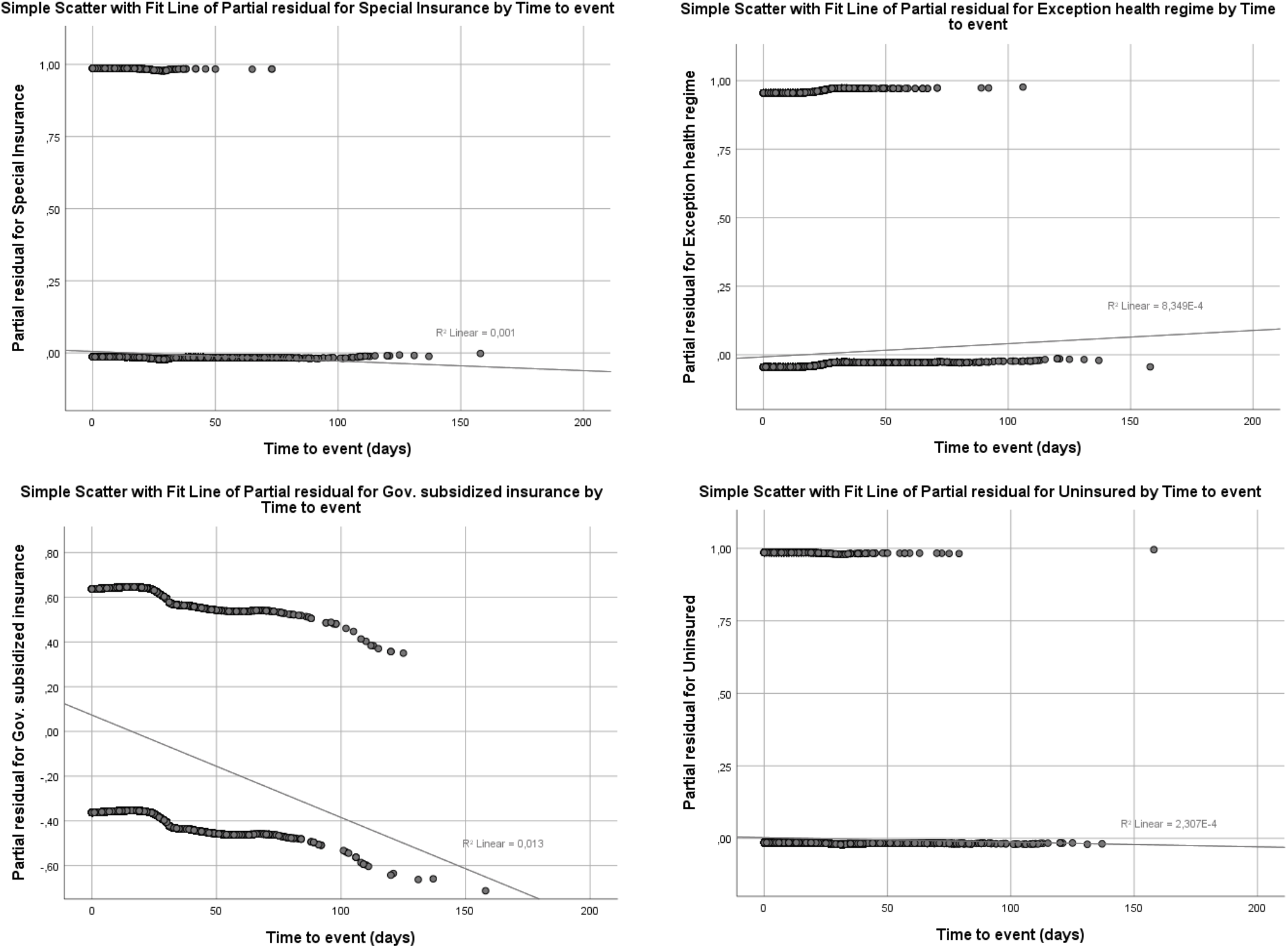
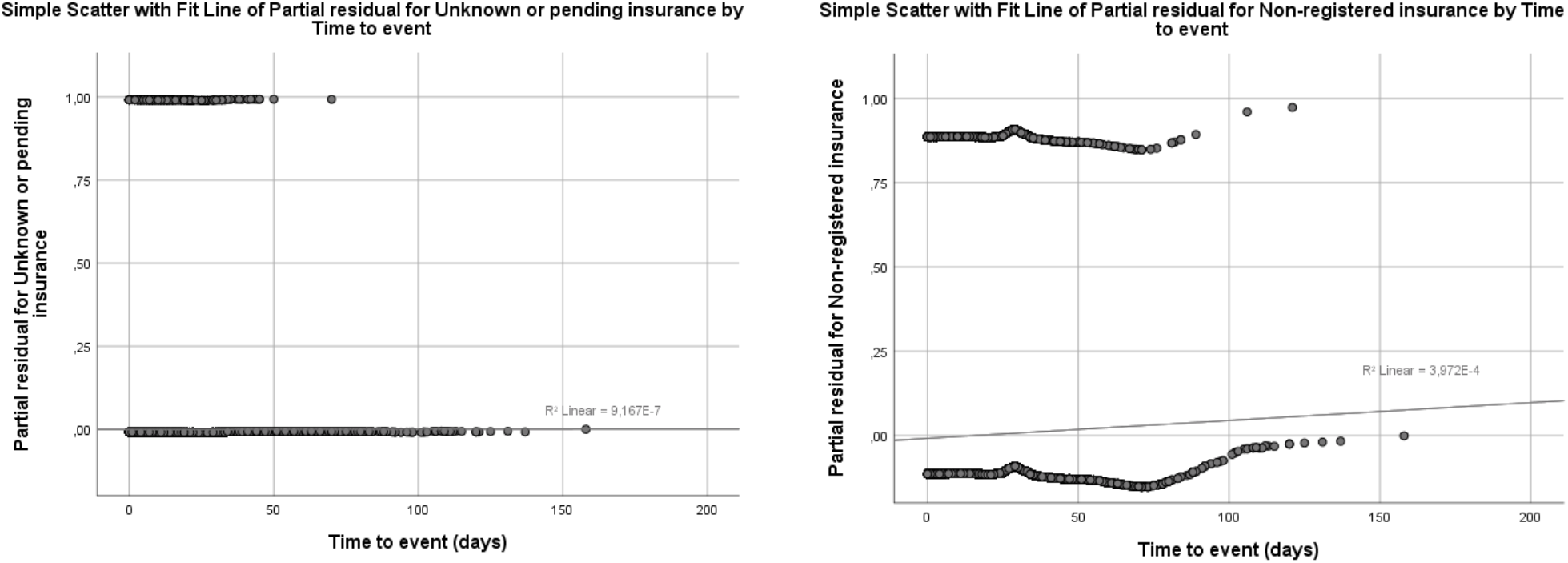
Partial residual vs time to event plots for Health insurance.

### Multi-predictor Cox Regression model with terms to address time-dependent factors

#### Numbers at risk

To complement the numbers at risk on figures of Survival plots for each predictor in the model, we present the number of the two types of censored cases for 0, 20, 40, 60, 80, 100, 120, 140, 160, 180 and 200 days of follow-up.

**Table S5.**
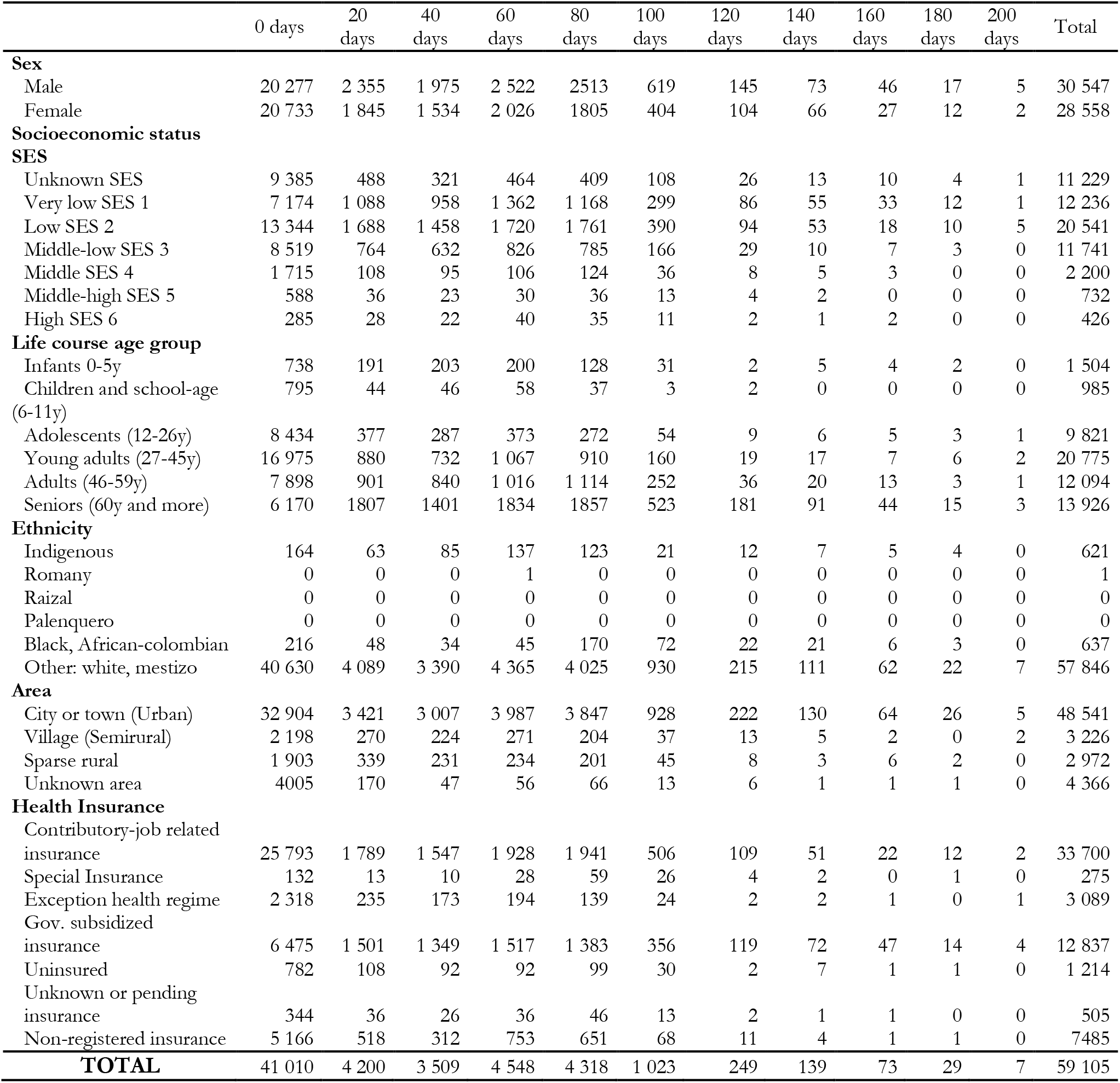
Censored symptomatic cases by follow-up time.

**Table S6.** Censored asymptomatic cases by follow-up time.

|  | 0 days | 20 days | 40 days | 60 days | 80 days | 100 days | 120 days | 140 days | 160 days | 180 days | 200 days | Total |
| --- | --- | --- | --- | --- | --- | --- | --- | --- | --- | --- | --- | --- |
| **Sex** |  |  |  |  |  |  |  |  |  |  |  |  |
| Male | 4 887 | 104 | 67 | 73 | 27 | 19 | 6 | 0 | 3 | 1 | 1 | 5 188 |
| Female | 5 279 | 73 | 47 | 46 | 11 | 9 | 8 | 3 | 1 | 1 | 0 | 5 478 |
| **Socioeconomic status** |  |  |  |  |  |  |  |  |  |  |  |  |
| **SES** |  |  |  |  |  |  |  |  |  |  |  |  |
| Unknown SES | 5 531 | 26 | 13 | 14 | 4 | 1 | 3 | 1 | 0 | 0 | 1 | 5 594 |
| Very low SES 1 | 469 | 21 | 11 | 13 | 3 | 2 | 4 | 0 | 3 | 1 | 0 | 527 |
| Low SES 2 | 1 754 | 63 | 50 | 49 | 21 | 13 | 4 | 2 | 1 | 0 | 0 | 1 957 |
| Middle-low SES 3 | 1 764 | 41 | 32 | 35 | 7 | 10 | 2 | 0 | 0 | 1 | 0 | 1 892 |
| Middle SES 4 | 418 | 12 | 4 | 6 | 1 | 2 | 1 | 0 | 0 | 0 | 0 | 444 |
| Middle-high SES 5 | 148 | 10 | 4 | 1 | 1 | 0 | 0 | 0 | 0 | 0 | 0 | 164 |
| High SES 6 | 82 | 4 | 0 | 1 | 1 | 0 | 0 | 0 | 0 | 0 | 0 | 88 |
| **Life course age group** |  |  |  |  |  |  |  |  |  |  |  |  |
| Infants 0-5y | 211 | 8 | 3 | 1 | 0 | 0 | 0 | 0 | 1 | 0 | 0 | 224 |
| Children and school-age (6-11y) | 270 | 0 | 0 | 0 | 0 | 0 | 0 | 0 | 0 | 0 | 0 | 270 |
| Adolescents (12-26y) | 2 186 | 15 | 4 | 7 | 4 | 0 | 0 | 0 | 1 | 0 | 0 | 2 217 |
| Young adults (27-45y) | 4 180 | 31 | 23 | 26 | 4 | 1 | 1 | 0 | 2 | 2 | 0 | 4 270 |
| Adults (46-59y) | 1 916 | 43 | 32 | 30 | 8 | 8 | 2 | 0 | 0 | 0 | 0 | 2 039 |
| Seniors (60y and more) | 1 403 | 80 | 52 | 55 | 22 | 19 | 11 | 3 | 0 | 0 | 1 | 1 646 |
| **Ethnicity** |  |  |  |  |  |  |  |  |  |  |  |  |
| Indigenous | 35 | 0 | 0 | 0 | 0 | 0 | 1 | 1 | 0 | 0 | 0 | 37 |
| Romany | 0 | 0 | 0 | 0 | 0 | 0 | 0 | 0 | 0 | 0 | 0 | 0 |
| Raizal | 0 | 0 | 0 | 0 | 0 | 0 | 0 | 0 | 0 | 0 | 0 | 0 |
| Palenquero | 0 | 0 | 0 | 0 | 0 | 0 | 0 | 0 | 0 | 0 | 0 | 0 |
| Black, African-colombian | 3 | 1 | 0 | 1 | 0 | 1 | 0 | 0 | 0 | 0 | 0 | 6 |
| Other: white, mestizo | 10 128 | 176 | 114 | 118 | 38 | 27 | 13 | 2 | 4 | 2 | 1 | 10 623 |
| **Area** |  |  |  |  |  |  |  |  |  |  |  |  |
| City or town (Urban) | 4 709 | 153 | 100 | 107 | 33 | 27 | 14 | 2 | 3 | 2 | 1 | 5 151 |
| Village (Semirural) | 72 | 0 | 3 | 1 | 2 | 0 | 0 | 1 | 0 | 0 | 0 | 79 |
| Sparse rural | 32 | 0 | 1 | 0 | 0 | 1 | 0 | 0 | 1 | 0 | 0 | 35 |
| Unknown area | 5 353 | 24 | 10 | 11 | 3 | 0 | 0 | 0 | 0 | 0 | 0 | 5 401 |
| **Health Insurance** |  |  |  |  |  |  |  |  |  |  |  |  |
| Contributory-job related insurance | 4 141 | 121 | 87 | 94 | 18 | 23 | 6 | 1 | 0 | 0 | 1 | 4 492 |
| Special Insurance | 14 | 1 | 0 | 0 | 0 | 0 | 0 | 0 | 2 | 1 | 0 | 18 |
| Exception health regime | 213 | 7 | 3 | 3 | 5 | 0 | 1 | 0 | 0 | 0 | 0 | 232 |
| Gov. subsidized insurance | 312 | 15 | 10 | 8 | 7 | 4 | 6 | 1 | 2 | 1 | 0 | 366 |
| Uninsured | 71 | 1 | 2 | 0 | 1 | 0 | 1 | 1 | 0 | 0 | 0 | 77 |
| Unknown or pending insurance | 9 | 0 | 1 | 0 | 1 | 0 | 0 | 0 | 0 | 0 | 0 | 11 |
| Non-registered insurance | 5 406 | 32 | 11 | 14 | 6 | 1 | 0 | 0 | 0 | 0 | 0 | 5 470 |
| TOTAL | 10 166 | 177 | 114 | 119 | 38 | 28 | 14 | 3 | 4 | 2 | 1 | 10 666 |

